# Is the Public Ready for a Tobacco-Free Ireland? A National Survey of Public Knowledge and Attitudes to Tobacco Endgame in Ireland

**DOI:** 10.1101/2022.12.01.22282993

**Authors:** Ellen Cosgrave, Martina Blake, Edward Murphy, Aishling Sheridan, Frank Doyle, Paul Kavanagh

**Affiliations:** Department of Public Health, Health Service Executive South East, Lacken, Dublin Road, Co. Kilkenny, Ireland R95 NV08; Health Service Executive (HSE) Tobacco-Free Ireland Programme, Health Service Executive, Dublin, Ireland; HSE Tobacco-Free Ireland Programme, Health Service Executive, Dublin, Ireland; Department of Health Psychology, School of Population Health, Royal College of Surgeons in Ireland, Dublin, Ireland; Department of Public Health and Epidemiology, School of Population Health, Royal College of Surgeons in Ireland, Dublin, Ireland

## Abstract

Tobacco-Free Ireland (TFI) policy sets a tobacco endgame goal to reduce smoking prevalence to less than 5% by 2025. However, public opinion on this goal, an important policy lever, is uncharted in Ireland. This study aimed to inform policy planning by measuring public knowledge and attitudes to tobacco endgame.

A telephone-administered cross-sectional survey of 1,000 randomly-dialled members of the general public was conducted. Prevalence of awareness, perceived achievability, and support for the TFI goal and potential tobacco endgame measures was calculated and compared across tobacco/e-cigarette user status. Logistic regression identified factors independently associated with TFI goal support.

Although TFI goal awareness was low (34.0%), support was high (74.6%), albeit most (60.2%) believed it achievable beyond 2025. Goal support was higher among non-tobacco/e-cigarette users (adjusted Odds Ratio (aOR) 2.68, 95%CI 1.83-3.90), women (aOR 1.55, 95%CI 1.13-2.14) and higher social class members (aOR 1.48, 95%CI 1.03-2.12). Product-focused measures were popular while views on user-focused measures were mixed: e.g. 86.1% supported nicotine content reduction while 40.3% supported user licensing. Phasing-out tobacco sales was highly-supported (82.8%); however, for most, this was contingent on support for currently addicted users.

Despite low awareness, there is strong support for tobacco endgame in Ireland and views on achievability are more realistic than the current goal of 2025. Supporting currently addicted tobacco users and engaging groups with lower support will be important policy planning and communication considerations. The Irish public are ready for tobacco endgame. These findings should re-invigorate policy planning to translate endgame ambition into action.

## Introduction

Ireland has a strong track-record in tobacco control.^1^ Through early adoption and progressive implementation of a high-impact policies,^2-4^ Ireland now ranks highest on the International Tobacco Control Scale.^5,6^ Smoking prevalence has almost halved in the last 20 years from 31% in 1998 to 17% in 2019.^7,8^ However, an increase in smoking prevalence to 18% in 2021 and a widening gap in smoking inequalities mean there is no room for complacency about the sufficiency of existing policies.^2,7^

Recognising a need to re-focus efforts from simply controlling tobacco-related harm to eliminating it completely, an ambitious shift in global policy discourse from “business-as-usual” tobacco control to “tobacco endgame” has emerged, which reframes solutions from a question of individual-level responsibility to a need for government-led action against a systemic societal issue.^9,10^ Tobacco endgame envisages a tobacco-free future involving policies, plans and interventions to end the tobacco epidemic.^10^ Fundamentally, it seeks to “change permanently the structural, political and social dynamics that sustain the tobacco epidemic, in order to end it within a specific time”.^10^ Potential tobacco endgame measures have been categorised under four themes:^11^ product-focused, targeting tobacco product appeal and addictiveness; institutional structure-focused, targeting tobacco industry; user-focused, targeting tobacco product affordability and access; supply-focused, targeting tobacco product availability and retailers.

As some countries enter the later stages of the tobacco epidemic,^12^ a number have set tobacco endgame goals. Through its current Tobacco-Free Ireland (TFI) policy, the Irish government declared a goal of reducing smoking prevalence to less than 5% by 2025.^13,14^ As of 2022, current smoking prevalence means this goal is unlikely to be met. Furthermore, the most recently announced legislative plan, which will provide for overdue protection of children from the sale of electronic cigarettes (e-cigarettes) (and related nicotine inhaling products) through a modern system of licencing for the tobacco and nicotine inhaling products retail sector,^15^ while welcome, lacks the “policy audacity” that will characterise tobacco endgame and which is already emerging in, for example, New Zealand.^16,17^ An alliance of local non-government organisations is advocating to augment these current legislative proposals to orient them more towards tobacco endgame.^18,19^ However, to-date political leaders have been slow to consider the ambitious measures demanded by their own TFI policy goal. This means Ireland, one of the first countries globally to adopt a tobacco endgame policy, is now likely to be the first country to fail to meet its own target.

Public support is a key lever for realising policy change.^16,20-23^ It creates a low risk political environment for policymakers, which is especially important for tobacco endgame where ideas are at the frontier of what seems imaginable.^16,24^

Although internationally a number of studies have examined public support for tobacco endgame goals and component potential measures, prior Irish studies have examined a limited number of endgame measures only and no study has explicitly assessed Irish public support for the current TFI goal.^25,26,27,28,29^ International studies have found high public support for tobacco endgame.^30,31,32^ Product and institutional structure-focused measures are especially well-supported, particularly reducing nicotine content in tobacco products,^33-37^ as well as requiring the tobacco industry to pay for costs caused by tobacco-related harm.^30,38-41^ However, support for user and supply-focused measures has been more varied. Although banning tobacco product sales near schools,^31,38-40,42,43^ reducing tobacco retailers in terms of place and number^31,38-40^ and increased taxation^24,27,40^ have been highly supported among the general population and “Tobacco 21” highly-supported by smokers,^33,34,42,44^ support for a tobacco sales phase-out over time has been found to be variable.^31,35,41,45-48^

Given the risk that the TFI goal may not be achieved by the current target of 2025, and recognising the role that public opinion could play in renewing policy planning, this study aimed to assess public opinion on tobacco endgame and component measures in Ireland so as to inform policy planning. Specific objectives were to measure public knowledge and attitudes to tobacco endgame in Ireland, to compare public knowledge and attitudes to tobacco endgame by tobacco/e-cigarette use status and to assess factors associated with TFI goal support.

## Materials and methods

A cross-sectional study was conducted to measure prevalence of public knowledge and attitudes to tobacco endgame and component endgame measures using a nationally representative sample of Irish adults. Literature review informed survey instrument development, identifying a set of endgame measures and sample questions to test support for them.^20,24-27,33,34,38-41,44,49,50^ These endgame measures and candidate questions were tested and refined through national and international expert consultation (See supplementary file).

### Sampling, recruitment and fieldwork

Sampling, recruitment, and data collection were conducted by an Irish-based market research company (IPSOS MRBI) from February 15^th^ to 28^th^ 2022. The target population was defined as members of the Irish general public aged 15 years or older including tobacco/e-cigarette users and non-users. Based on previous similar research in New Zealand it was hypothesised that prevalence of support for TFI may lie between 50-75%.^33^ A precision-based sample size was calculated based on the conservative assumption that 50% of the public reported support for TFI; a sample size of 784 was sufficient to measure this proportion with a 95% confidence interval (CI) of +/-3.5%; therefore a sample of size 1,000 was deemed more than sufficient for the purpose of the study.^51^

The market research company recruited a sample of 1,000 participants via random-digit-dialling using records of known mobile and landline prefixes provided by the Commission for Communications Regulation. The mobile:landline sample ratio was 85:15 and population coverage was estimated to be in excess of 99%. Quota sampling to reflect age, gender, region and social class enabled minor post-hoc weighting to adjust for generalisability where required. To obtain 1,000 complete responses, 3,386 individuals were contacted, equating to a response rate of 29.5% of those contacted. Participants were excluded if they were non-fluent in English or if they did not complete the survey in its entirety.

The survey consisted of 35 questions delivered via computer-aided telephone interviewing (CATI) as part of an omnibus survey (See questionnaire, supplementary file).^52^ Prior to administration, trained field interviewers were briefed on survey content including definitions of key terms and piloting was conducted. Informed verbal consent to participate was sought and secured prior to interviewing by trained interviewers. Questions were rotated and average completion time was 24 minutes. Fieldwork quality assurance procedures and data checking post-collection optimised data accuracy. Data were inputted via pre-coded responses (drop-down menus). To monitor interview quality, test interviews were monitored by supervisory staff. Ethical approval was granted by the Royal College of Physicians of Ireland Research Ethics Committee.

### Measures

Independent variables included demographic characteristics and tobacco/e-cigarette use behaviours (See data dictionary, supplementary file). To allow comparisons of key views among those at highest risk of tobacco-related harm, two variables (smoking status and e-cigarette use) were combined to produce the binary composite variable (tobacco/e-cigarette use); for this variable, those who responded ‘don’t know’ (n=6) were excluded from comparative analysis.

The principle dependent variables were support for the TFI goal and component tobacco endgame measures, which were dichotomised (‘support’/’no support’). ‘Support’ in this context was defined as agreement (‘strongly agree’/’somewhat agree’); ‘no support’ was defined as absence of support for such measures – aligning with previous literature this included those who responded ‘neither agree nor disagree’/’somewhat disagree’/’strongly disagree’/’don’t know’.^24 34 44^ Binary variables were also derived for views on government and Health Service Executive (HSE) action to tackle tobacco-related harm and for opinions on perceived TFI goal achievability.

### Analysis

Data were analysed using IBM SPSS Statistics Version 26.0.^53^ Frequency-based weights were applied throughout all analyses. Descriptive statistics were used determine counts and proportions of categorical demographic variables. Prevalence of public knowledge and attitudes to the TFI goal and views on government and HSE action to combat tobacco-related harm are presented as weighted prevalence estimates with 95% confidence intervals. Pearson’s Chi-squared test was used to compare differences in responses between tobacco/e-cigarette users and non-users. Prevalence of support for tobacco endgame measures was analysed among the total sample and sub-analysed by participant tobacco/e-cigarette use. Participant views regarding underlying pre-conditions and an acceptable timeframe for a tobacco sales phase-out were also analysed. Univariate logistic regression was used to assess associations between TFI goal support and respondent characteristics. Multivariable logistic regression modelling was then used to assess factors associated with TFI goal support, adjusting for relevant confounders.^31-33,47^ Confidence intervals were calculated for unadjusted and regression-adjusted odds ratios. Statistical significance was determined at the 0.05 level.

## Results

A total of 1,000 adults completed the survey in February 2022. Weighted sample characteristics for the overall sample are provided in Table 1. One-fifth (19.3%) were current tobacco/e-cigarette users; of these 57.3% were smokers only, 29.7% were e-cigarette users only and 13.0% were dual-users.

**Table 1:** Participant Characteristics.

| Variable | Valid Denominator | Total |  | Comparative Population Estimate |
| --- | --- | --- | --- | --- |
|  |  | N | % |  |
| **Gender** | 1,000 |  |  |  |
| Male |  | 491 | 49.1 | 48.8<sup>a</sup> |
| Female |  | 509 | 50.9 | 51.2<sup>a</sup> |
| **Age (years)** | 1,000 |  |  |  |
| 15-24 |  | 159 | 15.9 | 15.8<sup>b</sup> |
| 25-44 |  | 347 | 34.7 | 34.5<sup>b</sup> |
| 45-54 |  | 311 | 31.1 | 31.2<sup>b</sup> |
| 65+ |  | 183 | 18.3 | 18.5<sup>b</sup> |
| **Region** | 1,000 |  |  |  |
| Leinster |  | 558 | 55.8 | 55.3<sup>c</sup> |
| Munster |  | 267 | 26.7 | 26.9<sup>c</sup> |
| Connaught/Ulster |  | 175 | 17.5 | 17.8<sup>c</sup> |
| **Social class** | 1,000 |  |  |  |
| Higher (A,B,C1) |  | 435 | 43.5 | 43.5<sup>d</sup> |
| Lower (C2,D,E) |  | 505 | 50.5 | 50.5<sup>d</sup> |
| Farmer (F) |  | 60 | 6.0 | 6.0<sup>d</sup> |
| **Educational attainment\*\*** | 1,000 |  |  |  |
| Higher |  | 544 | 54.4 | 53<sup>e</sup> |
| Lower |  | 456 | 45.6 | 47<sup>e</sup> |
| **Smoker** | 999 |  |  |  |
| Yes |  | 137 | 13.7 | 18<sup>f</sup> |
| No |  | 862 | 86.3 | 82<sup>f</sup> |
| **E-cigarette user** | 995 |  |  |  |
| Yes |  | 83 | 8.3 | 4<sup>f</sup> |
| No |  | 912 | 91.7 | 96<sup>f</sup> |
| **Tobacco/e-cigarette user** | 994 |  |  |  |
| Yes |  | 192 | 19.3 | 20.3<sup>g</sup> |
| No |  | 802 | 80.7 | 79.7<sup>g</sup> |

\*Estimates for general adult population ≥15 years presented as available from sources;
\*\*Higher: had completed third level education, lower: had not completed third level education; Sources: **a:** Census, 2022;<sup>54</sup> **b:** Census, 2016;<sup>55</sup> **c:** Census, 2016;<sup>56</sup> **d:** Association of Irish Market Research Associations Estimates May 2020 (sourced from IPSOS MRBI); **e:** Labour Force Survey, 2021 (pertains to persons aged 25-64 only);<sup>57</sup> **f:** Healthy Ireland Survey, 2021;<sup>7</sup> **g:** Healthy Ireland Survey, 2019.<sup>58</sup>

### Views on Government Action to Tackle Tobacco-Related Harm

Most respondents (76.2%, 95% CI 73.6%-78.8%) believed the government should do more to tackle the harm done by smoking. There was a significant difference in this view between tobacco/e-cigarette users and non-users (68.4% versus 78.2, p=0.004). Less than half (42.2%, 95% CI 39.1%-45.3%) believed the government were doing enough to ensure the TFI goal is achieved.

### Awareness and Attitudes to the TFI Goal

One third of respondents (34.0%, 95% CI 31.1%-36.9%) were aware of the TFI goal. Three-quarters (74.6%, 95% CI 71.9%-77.3%) supported the TFI goal; a significant difference in support was evident between tobacco/e-cigarette users and non-users (54.4% versus 79.4%, p<0.001). The majority of respondents believed the TFI goal was achievable (76.6%, 95% CI 74.0%-79.2%); while 16.5% of respondents considered the goal achievable by 2025, most (60.2%) considered it achievable beyond 2025.

### Factors Associated with TFI Goal Support

Following adjustment, females (aOR 1.55, 95% CI 1.13-2.14, p=0.007), those of higher social class (aOR 1.48, 95% CI 1.03-2.12, p=0.035), and those of higher educational attainment (aOR 1.67, 95% CI 1.15-2.43, p=0.007) were significantly more likely to support the TFI goal than their comparative counterparts (Table 2). Compared to those aged 25-34 years, those aged 65 and older had almost 3 times higher odds of supporting the TFI goal (aOR 2.97, 95% CI 1.65-5.37, p<0.001) while non-tobacco/e-cigarette users remained significantly more likely to support the TFI goal compared to users (aOR 2.68, 95% CI 1.83-3.90, p<0.001).

**Table 2:** Multiple Logistic Regression Modelling Analysis of Participant Characteristics and TFI Goal Support (N=995)

| Characteristic | Unadjusted OR<br>(95% CI) | Adjusted OR<br>(95% CI) | P-value |
| --- | --- | --- | --- |
| **Gender** |  |  |  |
| Male | 1 | 1 |  |
| Female | **1.76 (1.31-2.35)** | **1.55 (1.13-2.14)** | **0.007** |
| **Age (years)** |  |  |  |
| 15-24 | 1.05 (0.64-1.71) | 1.59 (0.90-2.81) | 0.112 |
| 25-34 | 1 | 1 |  |
| 35-44 | 1.13 (0.71-1.82) | 1.34 (0.79-2.27) | 0.278 |
| 45-54 | 1.17 (0.72-1.92) | 1.24 (0.72-2.14) | 0.437 |
| 55-64 | 1.09 (0.65-1.82) | 1.68 (0.93-3.04) | 0.087 |
| ≥65 | **1.72 (1.03-2.86)** | **2.97 (1.65-5.37)** | **<0.001** |
| **Region** |  |  |  |
| Leinster | 1 | 1 |  |
| Munster | 1.08 (0.77-1.52) | 1.01 (0.69-1.47) | 0.975 |
| Connaught/Ulster | 1.02 (0.69-1.50) | 0.98 (0.64-1.52) | 0.938 |
| **Social class** |  |  |  |
| Lower (C2,D,E) | 1 | 1 |  |
| Higher (A,B,C1) | **1.94 (1.43-2.62)** | **1.48 (1.03-2.12)** | **0.035** |
| Farmer | **4.36 (1.83-10.42)** | **4.05 (1.63-10.10)** | **0.003** |
| **Educational attainment\*** |  |  |  |
| Lower | 1 | 1 |  |
| Higher | **1.73 (1.30-2.30)** | **1.67 (1.15-2.43)** | **0.007** |
| **Tobacco/e-cigarette user** |  |  |  |
| Yes | 1 | 1 |  |
| No | **3.22 (2.30-4.48)** | **2.68 (1.83-3.90)** | **<0.001** |
| **Prior awareness of the TFI goal** |  |  |  |
| Unaware | 1 | 1 |  |
| Aware | 1.11 (0.82-1.50) | 1.04 (0.74-1.46) | 0.829 |
| **Believed TFI goal was achievable** |  |  |  |
| No/Unsure | 1 | 1 |  |
| Yes | **4.03 (2.95-5.53)** | **4.90 (3.44-6.95)** | **<0.001** |

\*Higher: had completed third level education, lower: had not completed third level education; OR: odds ratio; CI: Confidence interval; Nagelkerke $r^2 = 0.232$ ; Percentage Accuracy in Classification = 74.6%; Bold font indicates $p < 0.05$ ; \*Adjusted for gender, age, region, social class, tobacco/e-cigarette use, prior TFI goal awareness and perceived achievability of the TFI goal.

### Support for Tobacco Endgame Measures

#### Product-Focused Measures

The highest prevalence of support was evident for lowering the nicotine content in tobacco products (86.1%, 95% CI 84.0%-88.2%) and e-cigarettes (85.6%, 95% CI 83.4%-87.8%) to make the products less addictive; and for tighter regulation of tobacco products (79.0%, 95% CI 76.5%-81.5%) (Figure 1). Compared to tobacco/e-cigarette users, non-users were significantly more likely to support all six product-focused measures (Supplementary Table 1).

**Figure 1:**
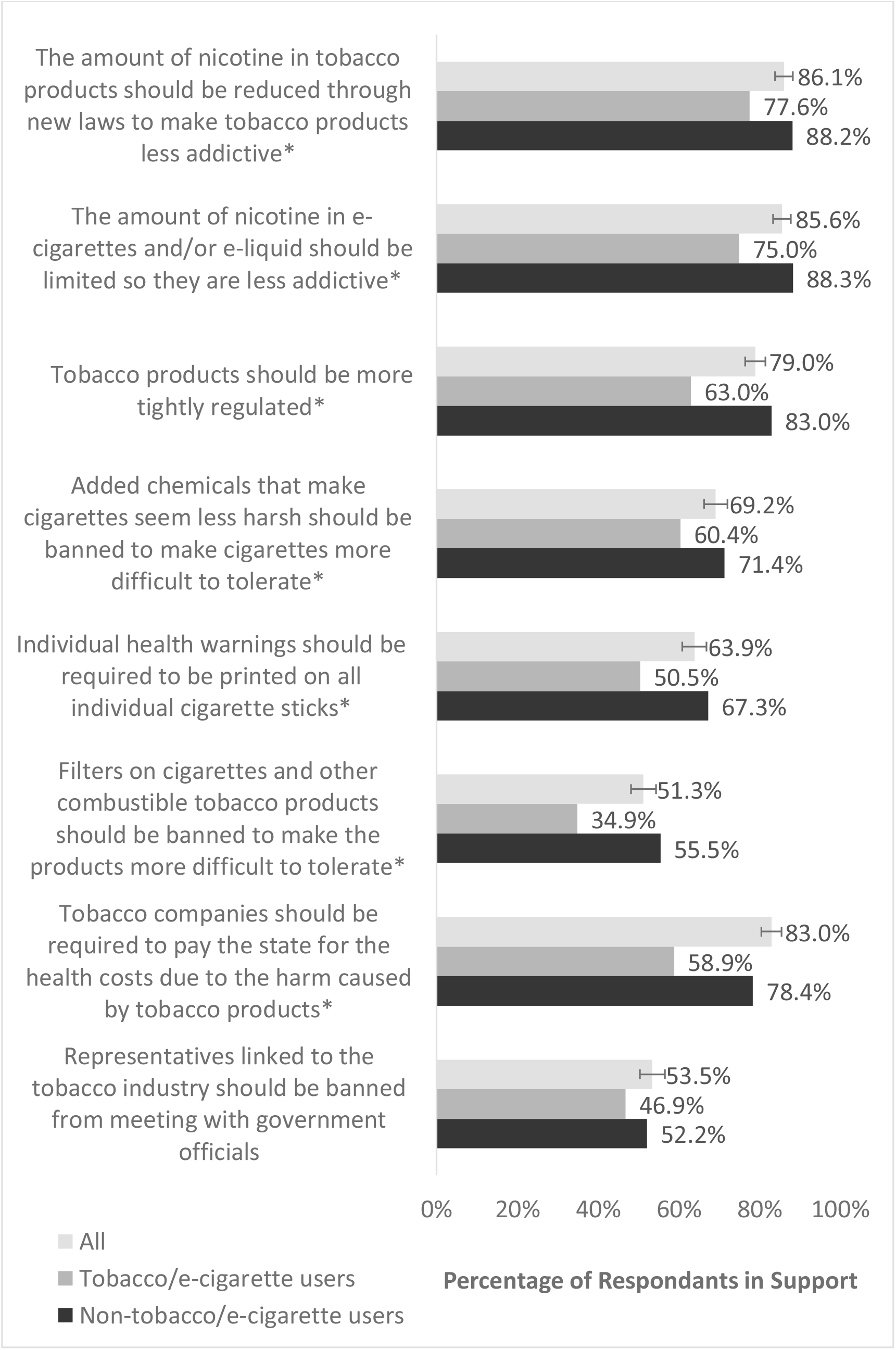
Percentage of Respondents who Supported Product-Focused and Institutional Structure-Focused Tobacco Endgame Measures (N=1,000)* Results are weighted and may not sum to totals; Error bars represent 95% CI; *Statistically significant difference between groups on chi squared test.

#### Institutional Structure-Focused Measures

More than three-quarters of respondents (78.4%, 95% CI 75.9%-81.0%) supported requiring tobacco companies to pay the state for the health costs accrued due to tobacco-related harm (Figure 1). A majority of respondents (52.2%, 95% CI 49.1%-55.3%) supported a ban on representatives linked to the tobacco industry meeting with government officials. Compared to tobacco/e-cigarette users, non-users were significantly more likely to support the former measure (83.0% versus 58.9%, p<0.001).

#### User-Focused Measures

The highest support was found for banning tobacco product sales near playgrounds, schools and university campuses (78.2%, 95% CI 75.6%-80.8%) and raising the legal age of purchasing tobacco products to 21 years and older (70.6%, 95% CI 67.8%-73.4%) (Figure 2). Compared to tobacco/e-cigarette users, non-users were significantly more likely to support all six user-focused measures.

**Figure 2:**
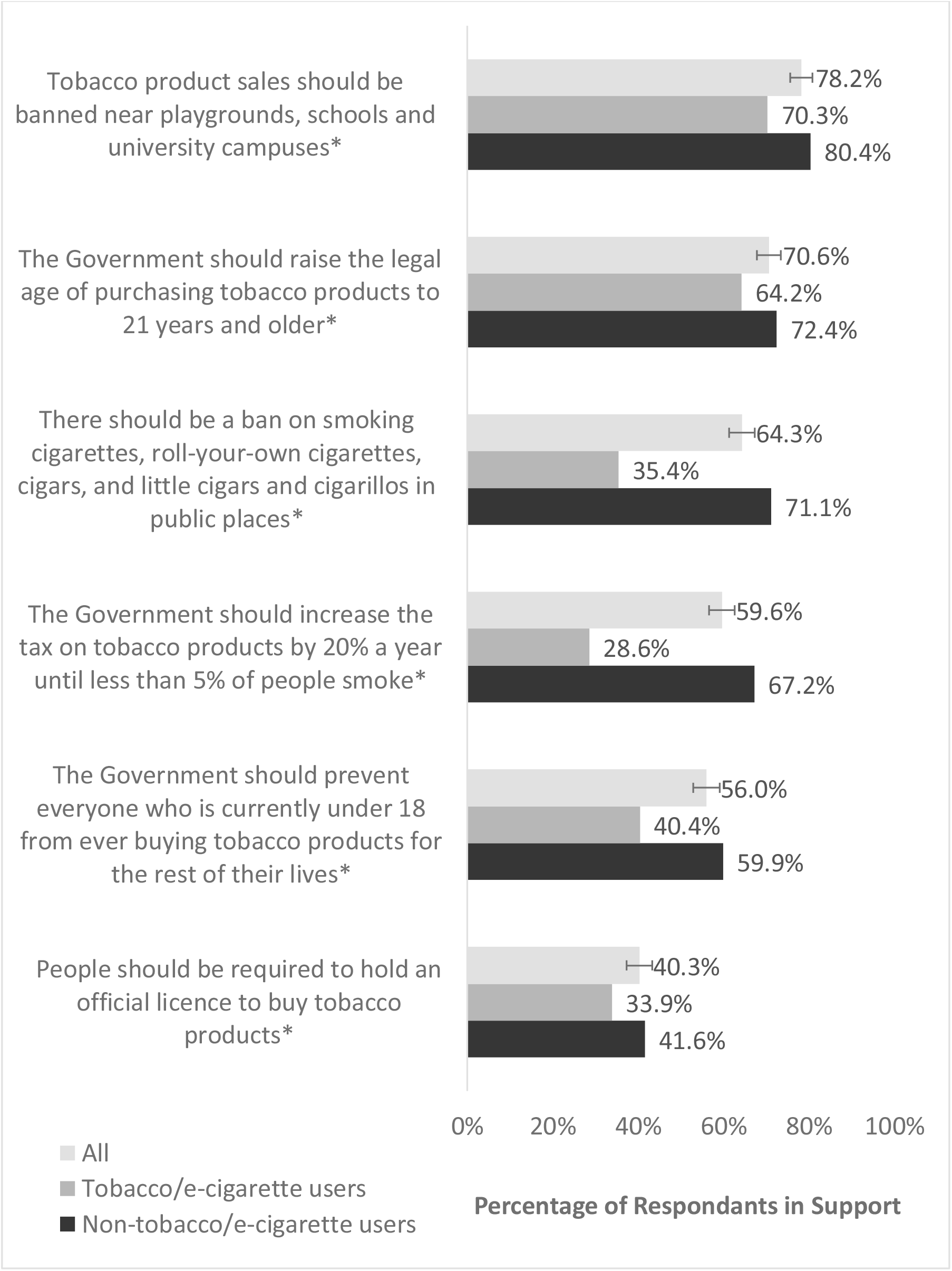
Percentage of Respondents who Supported User-Focused Tobacco Endgame Measures (N=1,000)* Results are weighted and may not sum to totals; Error bars represent 95% CI; *Statistically significant difference between groups on chi squared test.

#### Supply-Focused Measures

Highest support was evident for phasing out tobacco products sales (82.8%, 95% CI 80.5%-85.1%), and for requiring shops selling tobacco products to display information that encourages users to quit (81.9%, 95% CI 79.5%-84.3%) (Figure 3). Compared to tobacco/e-cigarette users, non-users where significantly more likely to support seven of eight supply-focused measures. Of those who supported a complete phase-out of tobacco product sales, for 85.0% support was contingent on provisions for people currently-addicted; either increased government assistance to help people who smoke to quit (74.8%) or allowing smokers to continue to buy tobacco products using a licence (40.8%). Views were similar across tobacco/e-cigarette use status. Two thirds (66.7%) of those who supported a complete phase-out believed this should occur within the next 10 years.

**Figure 3:**
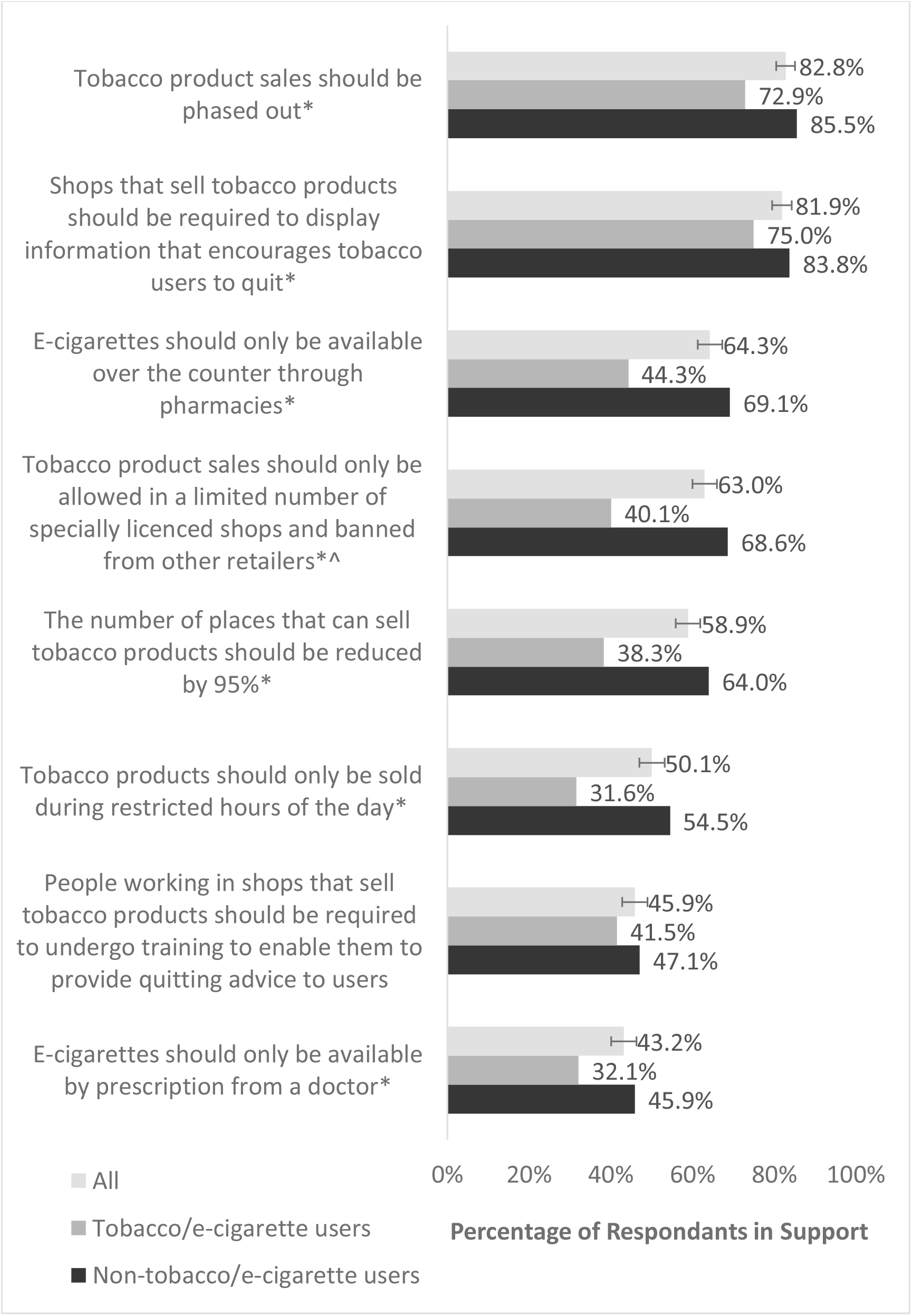
Percentage of Respondents who Supported Supply-Focused Tobacco Endgame Measures (N=1,000) Results are weighted and may not sum to totals; Error bars represent 95% CI; *Statistically significant difference between groups on chi squared test; ^Banned from smaller local shops, newsagents, off-licences and petrol stations.

## Discussion

### Key Findings

While Ireland was early to adopt a tobacco endgame goal through the TFI policy in 2013, achievement by the 2025 target is increasingly unlikely and current legislative plans lack the innovation and audacity needed to end the epidemic tobacco in the short-term.^59^ Public opinion has the potential to catalyse, shape and progress tobacco endgame.^11,16^ This study found that that despite low TFI goal awareness, 74.6% supported the goal and 76.6% believed it was achievable. Although lower among tobacco/e-cigarette users, goal support was still high at over 50%. There was majority public support for 19 of 22 endgame measures and particular support was evident for product-focused measures. A key finding was that high public support for phasing-out tobacco sales was contingent on supports for currently-addicted users. In addition to tobacco/e-cigarette use status, variation in public support was also evident across age, gender and socio-economic status.

### Knowledge and Attitudes to Tobacco Endgame in Ireland

High Irish public support for tobacco endgame among both tobacco/e-cigarette users and non-users was comparable to recent international findings.^30-32^ Such support was grounded in realism since a majority (54.3%) considered it achievable by 2035. Irish awareness of the TFI goal was lower than comparable evidence from New Zealand,^33^ suggesting scope to raise awareness. However, in this study, TFI goal awareness was not associated with support. Belief that government should do more to tackle smoking-related harm was higher than comparable findings from England and New Zealand, suggesting a mandate for policymakers to take action to progress endgame in Ireland.^30,33,47^

### Factors Associated with TFI Support

Consistent with international findings, females, non-tobacco/e-cigarette users and those aged 65 and older were more likely to support the TFI goal.^31^ Both higher educational attainment and higher social class were associated with increased goal support, indicating a positive association between goal support and socio-economic status. Against a context of widening inequalities in smoking in Ireland, this highlights the need for equitable endgame planning and communications. While belief the TFI goal was achievable was associated with increased goal support, contrary to previous literature, prior TFI goal awareness was not associated with increased support,^32^ suggesting support was high regardless of awareness.

### Support for Potential Tobacco Endgame Measures

All product-focused and institutional structure-focused measures had majority support. Particularly high support for reducing tobacco product nicotine content to make them less addictive aligned with international findings and policy commitments,^20,33-37,49,60,61^ most notably in New Zealand and the USA,^60,61^ as did requiring tobacco companies to pay for tobacco-related health costs.^30,38-41^ Support for a ban on filters was weaker, particularly among users. Tobacco industry misinformation may encourage belief that filters protect users – conversely, filters reduce the harshness of tobacco smoke, increasing toxic inhalant exposure.^62^

A majority of participants supported five of six user-focused measures and six of eight supply-focused measures. Mirroring national and most international literature, “Tobacco 21” was highly-supported by 70.6%,^28,30,38-42,63^ as were other youth-focused measures; e.g. banning sales near playgrounds, schools and universities.^31,38-40,42,43^ This finding is well-aligned with the priorities of non-governmental organisations in Ireland, which have already mobilised as an alliance to advocate for upward adjustment of age limits on tobacco retail.^18,19^ An exception was the Tobacco-Free Generation policy, which had less support, however support was still higher than most international literature.^33,38-40,45,64^ Question phrasing, which had prohibitionist connotations, may have influenced findings.^65^ Similarly, participants may have misunderstood unfamiliar endgame measures, suggesting the need to raise awareness of more complex proposals.

Support for a user-licence to purchase tobacco (40.3%) and for restricting e-cigarette sales to prescription-only access (43.2%) was lower than most international estimates.^38-40,45^ Substantial tax increases, a ban on tobacco use in public places and restricting tobacco retailers in terms of place, number and hours of sale, attracted minority support among tobacco/e-cigarette users. Support for raising tobacco taxation is likely to increase if revenue is used to fund healthcare initiatives,^33,42^ therefore it would be useful to assess if hypothecated tax increases would be more acceptable to users.

While the survey did not explore why certain measures were unpopular, some with less support shared a focus on individual-level restrictions for users, which could be perceived as punitive. Moreover, support for a phase out of tobacco sales was contingent on support for current users. Such findings indicate public opinion aligns with endgame principles which emphasise action tackling systems-factors perpetuating the tobacco epidemic over individual-level factors, and the importance of pursuing measures which support rather than punish current users.^10^

Support for a phase-out was higher than international findings,^31,35,41,45-48^ and much higher than in four Northern European countries including Ireland in 2012,^29^ suggesting support has increased in the past decade. This measure garnered higher support than less radical proposals e.g. restricting hours when tobacco can be sold. The term phase-out implies a gradual approach, potentially influencing findings.

Two-thirds of supporters believed a phase-out should occur within the next ten years –a higher proportion than international estimates, suggesting such a strategy could be acceptable in the medium-term.^46,66,67^ Congruent with international literature, support was highly-contingent upon measures for currently-addicted users.^24,33,49,50^ Requiring tobacco retailers to display information encouraging users to quit was also highly-supported, again emphasising the importance of quit supports. Conversely, congruent with findings from New Zealand, a majority of respondents did not support training retail staff to provide quitting advice at point-of-sale.^31^ Qualitative research, especially with current users, would be helpful to identify the most acceptable types of user supports.

Overall, support was significantly higher among non-tobacco/e-cigarette users for 20 of 22 endgame measures. Higher support among non-users for disruptive tobacco control measures is well-documented, particularly in Europe.^24,27,29,68^ However, evidence suggests support for novel measures can increase during and post-implementation, as was the case with the Irish workplace smoking ban and plain packaging in Australia.^6,69^ Therefore, comparatively lower support should not preclude implementation in the longer-term. However, further exploration of reasons for lower support for some measures is merited, particularly among users.

### Strengths and Limitations

This is the first study in Ireland providing timely and comprehensive data on knowledge and attitudes of tobacco endgame among the general Irish population. It fills an important research gap and brings the public voice into policy formulation on this key public health challenge.^70,71^ The cross-sectional study design was cost-efficient, time-efficient and appropriate for estimating prevalence of individual-level knowledge and attitudes.^72^ The large population-based sample afforded adequate statistical precision to estimate prevalence of key variables.

Notwithstanding measures to ensure sample representativeness, those not covered by the sampling frame (those without a registered mobile or landline number/without fluency in English) could not be included and no data were available on those who declined to participate; if such groups were systematically different to responders this could result in selection bias, specifically non-coverage and non-response bias respectively, reducing external validity. To mitigate such bias a post-hoc weighting strategy was employed. Still, comparisons with population estimates suggested smokers may have been under-represented and e-cigarette users over-represented, groups which had different views on the subject of the survey to the general population. However, weighted sample estimates for tobacco/e-cigarette use prevalence approximated corresponding population estimates.

Self-reported measurements may have introduced social desirability bias - stigma surrounding disagreement with tobacco control measures may have falsely overestimated true prevalence of support for endgame measures.^73^ Moreover, interviewer-administered telephone interviews are associated with increased risk of social desirability bias compared to self-administered surveys.^73^ Finally, although this study detected associations between TFI goal support and respondent characteristics, the cross-sectional nature of this study precludes causal inferences and omission of unmeasured variables from the final regression model (e.g. ethnicity) may have resulted in residual confounding.

## Conclusions

Global momentum on tobacco endgame is gathering, with some countries showing leadership by progressing fresh and ambitious measures. Ireland, a recognised tobacco control leader, risks being left behind. Through involving the public in discourse by assessing their knowledge and attitudes on tobacco endgame, this study should inform urgently required renewal of national endgame policy planning and should re-mobilise political commitment by evidencing public support for the bold actions needed to deliver on the TFI goal. A key finding was that public support for a tobacco sales phase-out was dependent on measures for current tobacco/e-cigarette users. Irish public interest that the pursuit of tobacco endgame is seen to support, not punish, current users is well aligned with the principles underpinning the concept and should be integral to endgame communication and planning. The range of endgame measures that were found to have high support, particularly product, supply and institutional structure-focused measures, provide a blueprint of priority actions (Supplementary Table 2) to achieve a Tobacco-Free Ireland. Some highly supported measures could easily be incorporated within the draft Public Health (Tobacco and Nicotine Inhaling Products) Bill due to enter the legislative process in the national parliament. The findings highlight a significant opportunity for political leadership in Ireland, which would benefit from public support, keep pace with international progress and deliver on the promise of ending the tobacco epidemic. While aspects of public opinion are inherently tied-up with specific national context, many of the findings in this study reinforce and develop evidence for international collaboration to translate endgame concepts into policy.

## Data Availability

All data produced in the present study are available upon reasonable request to the authors

## Statements

## Acknowledgements

The following colleagues gave input on the formulation and design of the survey: Prof Ruth Malone, University of California San Francisco Center for Tobacco Control Research and Education and Editor of Tobacco Control; Dr Elizabeth Smith and Dr Patricia McDaniel, University of California San Francisco Center for Tobacco Control Research and Education; Dr Rebecca Williams, Ms Elizabeth Anderson-Rodgers and Dr David Stupplebeen, California Tobacco Control Program, California Department of Public Health; Dr Fenton Howell, former National Tobacco Control Advisor, Department of Health; Ms Claire Gordon, Tobacco and Alcohol Control, Department of Health; Dr Helen McAvoy and Dr Ciara Reynolds, Institute of Public Health in Ireland; Dr Sara Burke, Centre for Health Policy and Management, Trinity College Dublin; Dr Daniela Rohde, Health Information and Quality Authority; Professor Des Cox and Members of the Royal College of Physicians of Ireland (RCPI) Tobacco Policy Group; and staff of the Department of Public Health, Health Service Executive South East.

High-level and summarised content of the study has been published as a policy brief and shared by the Health Service Executive Tobacco-Free Ireland Programme with key national stakeholders; in addition, the lead author has presented the findings orally at the European Public Health Meeting in 2022 (an abstract based on the conference proceedings were published here: Cosgrave, E., Blake, M., Murphy, E., Sheridan, A., Doyle, F., & Kavanagh, P. (2022). Is Ireland ready for tobacco endgame? A national survey of knowledge and attitudes to tobacco endgame: Ellen Cosgrave. The European Journal of Public Health, 32(Suppl 3), ckac129.034. https://doi.org/10.1093/eurpub/ckac129.034). The author was also invited to present at an online seminar organised by the Centre of Research Excellence on Achieving the Tobacco Endgame (https://tobacco-endgame.centre.uq.edu.au/event/session/780).

## Ethics approval

This study involved human participants and was approved by RCPI Ethics Committee (RECAP 157). Participants gave informed consent to participate in the study before taking part.

