## supplementary file for "Is the Public Ready for a Tobacco-Free Ireland? A National Survey of Public Knowledge and Attitudes to Tobacco Endgame in Ireland"

### Appendix A: Organogram of Stakeholder Consultations and Communications

Survey Instrument Development

Survey instrument feedback

HSE TFI Programme Team

HSE TFI Partners Group

RCPI Policy Group

USA

University of California San Francisco Center for Tobacco Control Research

California Tobacco Control Program, California Department of Public Health

Department of Health

Institute of Public Health

RCSI

TCD

HIQA

Fieldwork

Piloting

Department of Public Health South East

IPSOS MRBI

Delivery

IPSOS MRBI

*University of California San Francisco Center for Tobacco Control Research and Education and California Tobacco Control Program; **California Department of Public Health; HSE: Health Service Executive; TFI: Tobacco-Free Ireland; RCPI: Royal College of Physicians of Ireland; RCSI: Royal College of Surgeons in Ireland; TCD: Trinity College Dublin; HIQA: Health Information and Quality Authority.

### Appendix B: Survey Instrument

**Q.1.** Could I ask what age group you fall into?

| 15-17 | 1 |
| --- | --- |
| 18-24 | 2 |
| 25-34 | 3 |
| 35-44 | 4 |
| 45-54 | 5 |
| 55-64 | 6 |
| 65+ | 7 |

**Q.2.** Are you male or female?

| Male | 1 |
| --- | --- |
| Female | 2 |

**Q.3.** What region are you living in?

| Dublin | 1 |
| --- | --- |
| Rest of Leinster | 2 |
| Munster | 3 |
| Connaught/Ulster | 4 |

**Q.4.** To ensure we interview a wide cross-section of the public, could I first ask what the occupation of the chief income earner in your home is? (See glossary for definitions)

| Social Class A/B | 1 |
| --- | --- |
| Social Class C1 | 2 |
| Social Class C2 | 3 |
| Social Class D/E | 4 |
| Social Class F | 5 |

**Q.5.** What is the highest level of education you have completed to date?

| Completed primary school | 1 |
| --- | --- |
| Currently in secondary school | 2 |
| Completed secondary school | 3 |
| Currently at third level | 4 |
| Completed third level | 5 |
| No formal education | 6 |

**Q.6a** Do you smoke tobacco products? For the purposes of this survey, tobacco products do not include e-cigarettes. **READ OUT. SINGLE CODE**

| Yes | 1 |
| --- | --- |
| Yes, occasionaly | 2 |
| No | 3 |
| Don’t know | 4 |

**Q.6b** Which of the following statements BEST applies to you? **READ OUT. SINGLE CODE**

| I have never tried e-cigarettes | 1 |
| --- | --- |
| I have tried e-cigarettes but do not use them anymore | 2 |
| I have tried e-cigarettes and use them daily | 3 |
| I have tried e-cigarettes and still use them occasionally | 4 |
| Don’t know | 5 |

**Q.7a** Research shows that 18% of people aged 15 years and older in Ireland smoke. The Government of Ireland has a goal of becoming a tobacco-free country by 2025. This means reducing the proportion of Irish adults who smoke to less than 5%. Were you aware of this goal before now or were you not aware of this goal? **SINGLE CODE**

| Yes, aware | 1 |
| --- | --- |
| No, not aware | 2 |

**Q.7b** The ‘Tobacco-Free Ireland’ goal aims to reduce the proportion of Irish adults who smoke to less than 5% by 2025. Would you say you...... **READ OUT. SINGLE CODE. FLIP SCALE**

| Strongly support this goal | 1 |
| --- | --- |
| Support this goal | 2 |
| Are neutral about this goal | 3 |
| Oppose this goal | 4 |
| Strongly oppose this goal | 5 |
| Don’t know | 6 |

**Q.7c** Which of the following comes closest to your view - reducing smoking levels to less than 5% **READ OUT. SINGLE CODE. FLIP SCALE**

| Is achievable by 2025 | 1 |
| --- | --- |
| Is achievable but not until between 2026 and 2035 | 2 |
| Is achievable but not until between 2036 and 2050 | 3 |
| Is achievable but not until after 2050 | 4 |
| Is not achievable at all | 5 |
| Don’t know | 6 |

**Q.8a** Now I would like to talk about the role the government and the Health Service Executive (HSE) play in achieving Ireland’s Tobacco-Free Goal. To what extent do you agree or disagree with the following? **READ OUT. SINGLE CODE PER STATEMENT. ROTATE STATEMENTS. FLIP SCALE**

| **Sufficiency of national efforts** | **A** | **B** | **C** | **D** | **E** | **F** |
| --- | --- | --- | --- | --- | --- | --- |
| The Government should do more to tackle the harm done by smoking | 1 | 2 | 3 | 4 | 5 | 6 |
| The Government is doing enough to ensure that Ireland’s Tobacco-Free goal is achieved | 1 | 2 | 3 | 4 | 5 | 6 |
| The HSE is doing enough to tackle the harm done by smoking | 1 | 2 | 3 | 4 | 5 | 6 |

A: Strongly Agree; B: Somewhat agree; C: Neither Agree nor Disagree; D: Somewhat Disagree; E: Strongly Disagree; F: Don’t know

**Q.8b** Moving on, I am going to ask you about a number of potential measures that might help achieve the Tobacco-Free goal. To what extent do you agree or disagree with the following? **READ OUT. SINGLE CODE PER STATEMENT. ROTATE STATEMENTS. FLIP SCALE**

| **Views on Tobacco Endgame Measures** | **A** | **B** | **C** | **D** | **E** | **F** |
| --- | --- | --- | --- | --- | --- | --- |
| People should be required to hold an official licence to buy tobacco products | 1 | 2 | 3 | 4 | 5 | 6 |
| The number of places that can sell tobacco products should be reduced by 95% | 1 | 2 | 3 | 4 | 5 | 6 |
| Tobacco product sales should only be allowed in a limited number of specially licenced shops and banned from smaller local shops, newsagents, off-licences and petrol stations | 1 | 2 | 3 | 4 | 5 | 6 |
| Shops that sell tobacco products should be required to display information that encourages tobacco users to quit | 1 | 2 | 3 | 4 | 5 | 6 |
| People working in shops that sell tobacco products should be required to undergo training to enable them to provide quitting advice to tobacco users | 1 | 2 | 3 | 4 | 5 | 6 |
| The government should raise the legal age of purchasing tobacco products to 21 years and older | 1 | 2 | 3 | 4 | 5 | 6 |
| The government should prevent everyone who is currently under 18 from ever buying tobacco products for the rest of their lives | 1 | 2 | 3 | 4 | 5 | 6 |
| Tobacco product sales should be banned near playgrounds, schools and university campuses | 1 | 2 | 3 | 4 | 5 | 6 |
| There should be a ban on smoking cigarettes, roll-your-own cigarettes, cigars, and little cigars and cigarillos in public places | 1 | 2 | 3 | 4 | 5 | 6 |
| Tobacco products should only be sold during restricted hours of the day | 1 | 2 | 3 | 4 | 5 | 6 |
| The Government should increase the tax on tobacco products by 20% a year until less than 5% of people smoke | 1 | 2 | 3 | 4 | 5 | 6 |
| Tobacco products should be more tightly regulated | 1 | 2 | 3 | 4 | 5 | 6 |
| The amount of nicotine in tobacco products should be reduced through new laws to make tobacco products less addictive | 1 | 2 | 3 | 4 | 5 | 6 |
| Filters on cigarettes and other combustible tobacco products should be banned to make the products more difficult to tolerate | 1 | 2 | 3 | 4 | 5 | 6 |
| Added chemicals that make cigarettes seem less harsh should be banned to make cigarettes more difficult to tolerate | 1 | 2 | 3 | 4 | 5 | 6 |
| Individual health warnings should be required to be printed on all individual cigarette sticks | 1 | 2 | 3 | 4 | 5 | 6 |

A: Strongly Agree; B: Somewhat agree; C: Neither Agree nor Disagree; D: Somewhat Disagree; E: Strongly Disagree; F: Don’t know

**Q.8bi** Which of the following, if any, comes closest to your own view on the sale of tobacco products? **READ OUT. SINGLE CODE. ROTATE. FLIP CODES. INTERVIEWER INSTRUCTION: IF RESPONDENT ASKS, PHASING OUT OF TOBACCO SALES MEANS THE SALE OF TOBACCO PRODUCTS IN IRELAND WOULD BE GRADUALLY DISCONTINUED.**

| Tobacco product sales should be phased out | 1 |
| --- | --- |
| Tobacco product sales should be phased out but only if the government provides assistance to help smokers to quit | 2 |
| Tobacco product sales should be phased out but only if existing smokers can continue to buy tobacco products using a licence | 3 |
| Tobacco product sales should be phased out but only if the government provides assistance to help smokers to quit AND existing smokers can continue to buy tobacco products using a licence | 4 |
| Tobacco product sales should not be phased out | 5 |
| None of these/other option | 6 |
| Don’t know | 7 |

**Q.8bii** Over how many years do you think tobacco product sales should be phased out?	*_________years*

Over less than one year...................................................................98

Don’t know......................................................................................99

**Q.8c** Now I would like to discuss measures which target the tobacco industry. To

what extent do you agree or disagree with the following? **READ OUT. SINGLE CODE**

**PER STATEMENT. ROTATE STATEMENTS. FLIP SCALE**

| **Views on Industry Focused Tobacco-Endgame Measures** | **A** | **B** | **C** | **D** | **E** | **F** |
| --- | --- | --- | --- | --- | --- | --- |
| Tobacco companies should be required to pay the state for the health costs due to the harm caused by tobacco products | 1 | 2 | 3 | 4 | 5 | 6 |
| Representatives linked to the tobacco industry should be banned from meeting with government officials | 1 | 2 | 3 | 4 | 5 | 6 |

A=Strongly Agree; B=Somewhat agree; C=Neither Agree nor Disagree; D=Somewhat Disagree; E=Strongly Disagree; F=Don’t know

**Q.8d** Moving on to regulatory measures for e-cigarettes. To what extent do you agree or disagree with the following? **READ OUT. SINGLE CODE PER STATEMENT. ROTATE STATEMENTS. FLIP SCALE**

| **Views on E-cigarette Restrictions** | **A** | **B** | **C** | **D** | **E** | **F** |
| --- | --- | --- | --- | --- | --- | --- |
| E-cigarettes should only be available over the counter through pharmacies | 1 | 2 | 3 | 4 | 5 | 6 |
| E-cigarettes should only be available by prescription from a doctor | 1 | 2 | 3 | 4 | 5 | 6 |
| The amount of nicotine in e-cigarettes and/or e-liquid should be limited so they are less addictive | 1 | 2 | 3 | 4 | 5 | 6 |

A=Strongly Agree; B=Somewhat agree; C=Neither Agree nor Disagree; D=Somewhat Disagree; E=Strongly Disagree; F=Don’t know

### Appendix C: Data Dictionary

| No. | **Variable** | **Type of variable** | **Description** | **Original Coding** | **Recoding** |
| --- | --- | --- | --- | --- | --- |
| 1. | Age (years) | Categorical | Age group of participant | 1=15-17, 2=18-24, 3=25-34, 4=35-44, 5=45-54, 6=55-64, 7=65+ | 1=15-24 (1,2), 2=25-44 (3,4), 3=45-64 (5,6), 4=65+ (7) |
| 2. | Gender | Categorical | Gender of participant | 1=Male, 2=Female | 0=Female (2), 1=Male (1) |
| 3. | Region | Categorical | Region where participant resides | 1=Dublin, 2=Rest of Leinster, 3=Munster, 4=Connaught/Ulster | 0=Leinster (1,2), 1=Munster (3), 2=Connaught/Ulster (4) |
| 4. | Social class | Categorical | Social class of participant | 1=AB, 2=C1, 3=C2, 4=DE, 5=F | 0=Higher (1,2), 1=Lower (3,4),  2=Farmer (5) |
| 5. | Educational attainment | Categorical | Highest level of education attained | 1=Completed primary school, 2=Currently in secondary school, 3=Completed secondary school, 4=Currently at third level, 5=Completed third level, 6=No formal education | 0=Higher (5), 1=Lower (1-4,6) |
| 6. | Current smoking status | Categorical | Whether participant currently smokes cigarettes or tobacco products | 1=Yes daily smoker, 2=Yes occasionally, 3=No, 4=Don’t know | 0=Non-smoker (3),  1=Smoker (1,2),  Missing=Don’t know (4) |
| 7. | Current E-cigarette use status | Categorical | Whether participant currently uses e-cigarettes | 1=I have never tried e-cigarettes, 2=I have tried e-cigarettes but do not use them anymore, 3=I have tried e-cigarettes and use them daily, 4=I have tried e-cigarettes and use them occasionally, 5=Don’t know | 0=Non-e-cigarette user (1,2), 1=E-cigarette user (3,4), Missing=Don’t know (5) |
| 8. | Current tobacco/e-cigarette use status | Categorical | Whether participant currently uses tobacco products or e-cigarettes | Composite variable derived from variables 6. And 7. | 0=Non-tobacco/e-cigarette user, 1=Tobacco/e-cigarette user, Missing=Don’t know |
| 9. | TFI goal awareness | Categorical | Whether participant is aware of TFI goal | 1=Yes, aware, 2=No, not aware | 0=Not aware (2), 1=Aware (1) |
| 10. | TFI goal support | Categorical | Whether participant supports the TFI goal | 1=Strongly agree, 2=Somewhat agree, 3=Neither agree nor disagree, 4=Somewhat disagree, 5=Strongly disagree, 6=Don’t know | 0=No support (3,4,5,6), 1=Support (1,2) |
| 11. | TFI goal perceived achievability | Categorical | Whether participant believes the TFI goal is achievable | 1=Is achievable by 2025, 2=Is achievable but not until between 2026 and 2035, 3=Is achievable but not until between 2036 and 2050, 4=Is achievable but not until after 2050, 5=Is not achievable at all, 6=Don’t know | 0=Not achievable/Don’t know (5,6), 1=Achievable (1,2,3,4) |

**Appendix C (Continued)**

| No. | **Variable** | **Type of variable** | **Description** | **Original Coding** | **Recoding** |
| --- | --- | --- | --- | --- | --- |
| 12. | TFI goal achievability timeframe | Categorical | Timeframe by which participant believes the TFI goal is achievable | As above | 1=Is achievable by 2025, 2=Is achievable but beyond 2025, 3=Is not achievable/Don’t know |
| 13. | View on government action on smoking-related harm | Categorical | Whether participant agreed government should do more to tackle smoking-related harm | 1=Strongly agree, 2=Somewhat agree, 3=Neither agree nor disagree, 4=Somewhat disagree, 5=Strongly disagree, 6=Don’t know | 0=Do not agree/Don’t know (3,4,5,6), 1=Agree (1,2) |
| 14. | View on HSE action on smoking-related harm | Categorical | Whether participant agreed the HSE is doing enough to tackle smoking-related harm | 1=Strongly agree, 2=Somewhat agree, 3=Neither agree nor disagree, 4=Somewhat disagree, 5=Strongly disagree, 6=Don’t know | 0=Do not agree/Don’t know (3,4,5,6), 1=Agree (1,2) |
| 15. | View on government commitment to TFI goal | Categorical | Whether participant agreed Government is doing enough to ensure TFI is achieved | 1=Strongly agree, 2=Somewhat agree, 3=Neither agree nor disagree, 4=Somewhat disagree, 5=Strongly disagree, 6=Don’t know | 0=Do not agree/Don’t know (3,4,5,6), 1=Agree (1,2) |
| 16.-37. | Support for 21 component endgame measures (as outlined in Appendix F) | Categorical | Whether participant supported proposed endgame measures | 1=Strongly agree, 2=Somewhat agree, 3=Neither agree nor disagree, 4=Somewhat disagree, 5=Strongly disagree, 6=Don’t know | 0=No Support (3,4,5,6), 1=Support (1,2) |
| 38. | Support for a tobacco sales phase-out | Categorical | Whether participants supported a tobacco sales phase-out | 1=Support with no conditions, 2=Support if the government provides assistance to help smokers to quit, 3=Support but only if existing smokers can continue to buy tobacco products using a licence, 4=Support but only if conditions in both 2. and 3. are met , 5=Does not support , 6=Don’t know | 0=No support(5,6), 1=Support (1,2,3,4) |
| 39. | Acceptable phase-out timeline | Categorical | Timeframe within which participants supported a tobacco sales phase-out | Enter as given, 11=11 years or longer, 12=less than 1 year, 13=Don’t know | 1=0-5 years, 2=6-10 years, 3=>10 years, 4=Don’t know |

### Appendix D: Support for Tobacco Endgame Measures*

| **Measure** | **Total**  **n (%, 95% CI)** | ***Tobacco/e-cigarette User***  ***n (%)*** | ***Non-Tobacco/e-cigarette User***  ***n (%)*** | ***P-value*** |
| --- | --- | --- | --- | --- |
| ***Lowering the nicotine content in tobacco products*** | **(N=1,000)** | ***(N=182)*** | ***(N=802)*** |  |
| Support | 861 (86.1, 84.0-88.2) | *149 (77.6)* | *707 (88.2)* | *<0.001* |
| No Support | 139 (13.9, 11.8-16.0) | *43 (22.4)* | *95 (11.8)* |  |
| ***Lowering the nicotine content in e-cigarettes*** |  |  |  |  |
| Support | 856 (85.6, 83.4-87.8) | *144 (75.0)* | *708 (88.3)* | *<0.001* |
| No Support | 144 (14.4, 12.2-16.6) | *48 (25.0)* | *94 (11.7)* |  |
| ***Tighter regulation of tobacco products*** |  |  |  |  |
| Support | 790 (79.0, 76.5-81.5) | 121 (63.0) | 666 (83.0) | *<0.001* |
| No Support | 210 (21.0, 18.5-23.5) | 71 (37.0) | 136 (17.0) |  |
| ***Ban on added chemicals that make cigarettes seem less harsh*** |  |  |  |  |
| Support | 692 (69.2, 66.3-72.1) | *116 (60.4)* | *573 (71.4)* | *0.003* |
| No Support | 308 (30.8, 27.9-33.7) | *76 (39.6)* | *229 (28.6)* |  |
| ***Requiring individual health warnings on all individual cigarette sticks*** |  |  |  |  |
| Support | 639 (63.9, 60.9-66.9) | *97 (50.5)* | *540 (67.3)* | *<0.001* |
| No Support | 361 (36.1, 33.1-39.1) | *95 (49.5)* | *262 (32.7)* |  |
| ***Banning filters on cigarettes and other combustible tobacco products*** |  |  |  |  |
| Support | 513 (51.3, 48.2-54.4) | *67 (34.9)* | *445 (55.5)* | *<0.001* |
| No Support | 487 (48.7, 45.6-51.8) | *125 (65.1)* | *357 (44.5)* |  |
| ***Requiring tobacco companies to pay the state for the health costs due to tobacco-related harm*** |  |  |  |  |
| Support | 784 (78.4, 75.9-81.0) | *113 (58.9)* | *666 (83.0)* | *<0.001* |
| No Support | 216 (21.6, 19.1-24.2) | *79 (41.1)* | *136 (17.0)* |  |

**Appendix D (continued)**

| **Measure** | **Total**  **n (%, 95% CI)** | ***User***  ***n (%)*** | ***Non- User***  ***n (%)*** | ***P-value*** |
| --- | --- | --- | --- | --- |
| ***Banning tobacco industry representatives meeting with government*** | **(N=1,000)** | ***(N=182)*** | ***(N=802)*** |  |
| Support | 522 (52.2, 49.1-55.3) | *90 (46.9)* | *429 (53.5)* | *0.099* |
| No Support | 478 (47.8, 44.7-50.9) | *102 (53.1)* | *373 (46.5)* |  |
| ***Banning tobacco product sales near playgrounds, schools and universities*** |  |  |  |  |
| Support | 782 (78.2, 75.6-80.8) | *135 (70.3)* | *645 (80.4)* | *0.002* |
| No Support | 218 (21.8, 19.2-24.4 | *57 (29.7)* | *157 (19.6)* |  |
| ***“Tobacco 21” policy*** |  |  |  |  |
| Support | 706 (70.6, 67.8-73.4) | *124 (64.2)* | *581 (72.4)* | *0.024* |
| No Support | 294 (29.4, 26.6-32.2) | *69 (35.8)* | *221 (27.6)* |  |
| ***Ban on smoking tobacco products in public places*** |  |  |  |  |
| Support | 643 (64.3, 61.3-67.3) | *68 (35.4)* | *570 (71.1)* | *<0.001* |
| No Support | 357 (35.7, 32.7-38.7) | *124 (64.6)* | *232 (28.9)* |  |
| ***Tax increases of 20%+ per year until <5% of the population smoke*** |  |  |  |  |
| Support | 596 (59.6, 56.6-62.6) | *55 (28.6)* | *539 (67.2)* | *<0.001* |
| No Support | 404 (40.4, 37.4-43.4) | *137 (71.4)* | *263 (32.8)* |  |
| ***“Tobacco-Free Generation” policy*** |  |  |  |  |
| Support | 560 (56.0, 52.9-59.1) | *78 (40.4)* | *480 (59.9)* | *<0.001* |
| No Support | 440 (44.0, 40.9-47.1) | *115 (59.6)* | *322 (40.1)* |  |
| ***Tobacco user-licence*** |  |  |  |  |
| Support | 403 (40.3, 37.3-43.4) | *65 (33.9)* | *334 (41.6)* | *0.048* |
| No Support | 597 (59.7, 56.7-62.7) | *127 (66.1)* | *468 (58.4)* |  |
| ***Complete phase-out of tobacco product sales*** |  |  |  |  |
| Support | 828 (82.8, 80.5-85.1) | *140 (72.9)* | *686 (85.5)* | *<0.001* |
| No Support | 172 (17.2, 14.9-19.5) | *52 (27.1)* | *116 (14.5)* |  |

**Appendix D (continued)**

| **Measure** | **Total**  **n (%, 95% CI)** | ***User***  ***n (%)*** | ***Non-User***  ***n (%)*** | ***P-value*** |
| --- | --- | --- | --- | --- |
|  | **(N=1,000)** | ***(N=182)*** | ***(N=802)*** |  |
| ***Requiring tobacco retailers to display information encouraging users to quit*** |  |  |  |  |
| Support | 819 (81.9, 79.5-84.3) | *144 (75.0)* | *672 (83.8)* | *0.004* |
| No Support | 181 (18.1, 15.7-20.5) | *48 (25.0)* | *130 (16.2)* |  |
| ***Restricting e-cigarette sales to over the counter sales in pharmacies*** |  |  |  |  |
| Support | 643 (64.3, 61.3-67.3) | *85 (44.3)* | *554 (69.1)* | *<0.001* |
| No Support | 357 (35.7, 32.7-38.7) | *107 (55.7)* | *248 (30.9)* |  |
| ***Allowing tobacco sales in a limited number of specially licenced shops*** |  |  |  |  |
| Support | 630 (63.0, 60.0-66.0) | *77 (40.1)* | *550 (68.6)* | *<0.001* |
| No Support | 370 (37.0, 34.0-40.0) | *115 (59.9)* | *252 (31.4)* |  |
| ***Reducing the number of places that can sell tobacco products by 95%*** |  |  |  |  |
| Support | 589 (58.9, 55.9-62.0) | *74 (38.3)* | *513 (64.0)* | *<0.001* |
| No Support | 411 (41.1, 38.1-44.2) | *119 (61.7)* | *289 (36.0)* |  |
| ***Restricting tobacco product sales to restricted hours of the day*** |  |  |  |  |
| Support | 501 (50.1, 47.0-53.2) | *61 (31.6)* | *437 (54.5)* | *<0.001* |
| No Support | 499 (49.9, 46.8-53.0) | *132 (68.4)* | *365 (45.5)* |  |
| ***Requiring workers that sell tobacco to undergo training to provide quitting advice*** |  |  |  |  |
| Support | 459 (45.9, 42.8-49.0) | *80 (41.5)* | *377 (47.1)* | *0.160* |
| No Support | 541 (54.1, 51.0-57.2) | *113 (58.5)* | *424 (52.9)* |  |
| ***Restricting e-cigarette sales to prescription-only access*** |  |  |  |  |
| Support | 432 (43.2, 40.1-46.3) | *62 (32.1)* | *368 (45.9)* | *0.001* |
| No Support | 568 (56.8, 53.7-59.9) | *131 (67.9)* | *433 (54.1)* |  |

User: tobacco/e-cigarette user; Non-user: non-tobacco/e-cigarette user; *Results are weighted and may not sum to totals.

### Appendix E: Summary of Public Support Levels for Tobacco Endgame Measures

|  |  | Support* | | |
| --- | --- | --- | --- | --- |
| Category | Tobacco Endgame Measure | Total sample | Tobacco/e-cigarette users | Non-users |
| Product-Focused | Lowering the nicotine content in tobacco products | High | High | High |
|  | Lowering the nicotine content in e-cigarettes | High | High | High |
|  | Tighter regulation of tobacco products | High | Majority | High |
|  | Ban on added chemicals that make cigarettes seem less harsh | Majority | Majority | High |
|  | Requiring individual health warnings on all individual cigarette sticks | Majority | Majority | Majority |
|  | Banning filters on cigarettes and other combustible tobacco products | Majority | Low | Majority |
| Institutional Structure-Focused | Requiring tobacco companies to pay for tobacco-related health costs due to tobacco-related harm | High | Majority | High |
|  | Banning tobacco industry representatives meeting with government | Majority | Low | Majority |
| User-Focused | Banning tobacco product sales near playgrounds, schools and universities | High | High | High |
|  | “Tobacco 21” policy | High | Majority | High |
|  | Ban on smoking tobacco products in public places | Majority | Low | High |
|  | Tax increases of 20%+ per year until <5% of the population smoke | Majority | Low | Majority |
|  | “Tobacco-Free Generation” policy | Majority | Low | Majority |
|  | Tobacco user-licence | Low | Low | Low |
| Supply-Focused | Complete phase-out of tobacco product sales | High | High | High |
|  | Requiring tobacco retailers to display information encouraging users to quit | High | High | High |
|  | Restricting e-cigarette sales to over the counter sales in pharmacies | Majority | Low | Majority |
|  | Allowing tobacco sales in a limited number of specially licenced shops | Majority | Low | Majority |
|  | Reducing the number of places that can sell tobacco products by 95% | Majority | Low | Majority |
|  | Restricting tobacco product sales to restricted hours of the day | Majority | Low | Majority |
|  | Requiring tobacco sales staff to undergo training to provide quitting advice | Low | Low | Low |
|  | Restricting e-cigarette sales to prescription-only access | Low | Low | Low |

*Levels of support are defined as follows: High = ≥70%; Majority = >50% - <70%; Low = 0-50%.
